# Analysis and Estimation of Length of In-Hospital Stay Using Demographic Data of COVID-19 Recovered Patients in Singapore

**DOI:** 10.1101/2020.04.17.20069724

**Authors:** Jaya Sreevalsan-Nair, Reddy Rani Vangimalla, Pritesh Rajesh Ghogale

**Affiliations:** Graphics-Visualization-Computing Lab *and* E-Health Research Center, International Institute of Information Technology Bangalore, Karnataka 560100, India

**Keywords:** COVID-19, Length of in-hospital stay, Descriptive statistics, Hypothesis testing, Regression

## Abstract

Singapore is one of the countries which has taken early, systematic, and rigorous nationwide responses to slowing the COVID-19 contagion down. By February 4, 2020, the government of Singapore restricted the mobility of the vulnerable age-groups. In the current study, we study the influence of the age-based vulnerability of the population with respect to COVID-19 conditions on the recovery of COVID-19 patients. We study 245 patients in Singapore recovered and discharged during January 23–April 01. We first study the descriptive statistics of the length of in-hospital stay (LOS) of the COVID-19 patients based on demographic variables, namely age, and gender. Then, we determine the distribution of LOS, using local and generalized linear regression models. We take the approach of periodization based on critical changes in the disease transmission model. Even though the overall recovery rate has reduced drastically after a sudden spike in daily confirmations, our analysis shows that there is a considerable shift in the COVID-19 confirmations to the population in the non-vulnerable age-groups. We show that the LOS of the non-vulnerable age group is considerably lower at 9 days, as opposed to 15 or 20 days in the existing literature.

## 1 Introduction

The viral contagion, named COVID-19, was declared as a global pandemic by the World Health Organization (WHO) on March 11, 2020.^2^ The pandemic, characterized by atypical pneumonia, is caused by a virus from the coronavirus family, namely SARS-CoV2 (Severe Acute Respiratory Syndrome Coronavirus-2), which is a positive-sense single-stranded RNA virus. As of April 2, there were 827,419 positively confirmed cases, and 40,777 deaths, spread across 206 countries^3^, and the ratio of the number of the recovered^4^ to the infected patients, *r_ri_* was ~0.2. On the other hand, in Singapore, 245 of 1000 COVID-19 positive cases had clinically recovered by April 1, giving *r_ri_*=0.245. Aside from the recovered/hospital-discharged cases, the first two deaths in Singapore owing to the COVID-19 complications were only three by April 1. The count of the recovered was ~79.5% of closed cases (i.e., recovery and death) worldwide, whereas ~98.8% in Singapore. Another Southeast Asian country with similar COVID-19 response, South Korea crossed its first 1000 COVID-19 confirmations^5^ during January 20-February 26, i.e., within 38 days, compared to 70 days in Singapore, giving *r_ri_* = 0.019 with the recovered being only 66.67% of closed cases.

Singapore has fared well owing to its effective public healthcare system that implemented a quick and efficient pandemic response since its first case got confirmed on January 23. The government enforced strict quarantine, isolation, hospital surveillance, large-scale contact tracing, and testing. The centralization of the system has facilitated responsive data gathering and anonymized patient-wise reporting to the public. The first cases of local transmission were determined on February 4, after which there were concerted efforts to protect the age-groups, namely, (0-20) and (60+) years, considered vulnerable to the COVID-19 conditions. South Korea has implemented similar measures but, did not focus on the early containment of local transmission, unlike Singapore. Thus, the overall slow transmission of COVID-19 in Singapore indicates that its early measures have been effectively slowed local transmission.

In this study, we focus on the influence of the age-based vulnerability of the population on the clinical recovery of the patients in Singapore during January 23-April 1. We hypothesize that these containment measures have indirectly reduced the length of in-hospital stay (LOS), Δ*t*_LOS_, for COVlD-19. We use the open data available through the official press releases of the Ministry of Health (MoH), Government of Singapore, for this study. Thus, we emphasize on the credibility of our study owing to the *reliability, accessibility*, and *availability* of the data [1]. A demographic analysis of the discharged patients complements similar demographic-related observations in differential disease transmission, e.g., higher fatality in older males with comorbidities [2, 3]. We perform piecewise analyses for estimating time-varying Δ*t*_LOS_ using periodization, similar to the estimation of reproduction number [4, 5]. We study the temporal changes in the age-gender distribution of the COVID-19 patients and discharged ones to estimate Δ*t*_LOS_. We then determine the statistical significance of the demographic variables and period on Δ*t*_LOS_, and find appropriate distributions of the LOS and fitting multivariate regression models. The novel contribution of our work is in linking the temporal reduction in Δ*t*_LOS_ to the demographic structure.

## 2 Methods

The data for our work has been collated from the case summaries in the public press releases made by the MoH^6^. We use the age, gender, positive confirmation date, and discharge date from these anonymized summaries. We define the *length of in-hospital stay* Δ*t*_LOS_ as the time elapsed between positive and two consecutive negative COVID-19 confirmations using the real-time reverse transcription polymerase chain reaction (RT-PCR) tests. Owing to the strict protocols followed by the Singapore healthcare system, the COVID-19 patients are hospitalized upon positive confirmation. Hence, recovery-period here is equivalent to the *virus shedding period*, which is estimated to be 15 days [6] or 20 days [7].Overall, we study the Δ*t*_LOS_ of 245 patients who were discharged by April 1 (§Figure 1). (*Additional data along with supporting details of the pandemic response in Singapore until April 1 are given in Section 1, Supplementary File*.)

**Figure 1:**
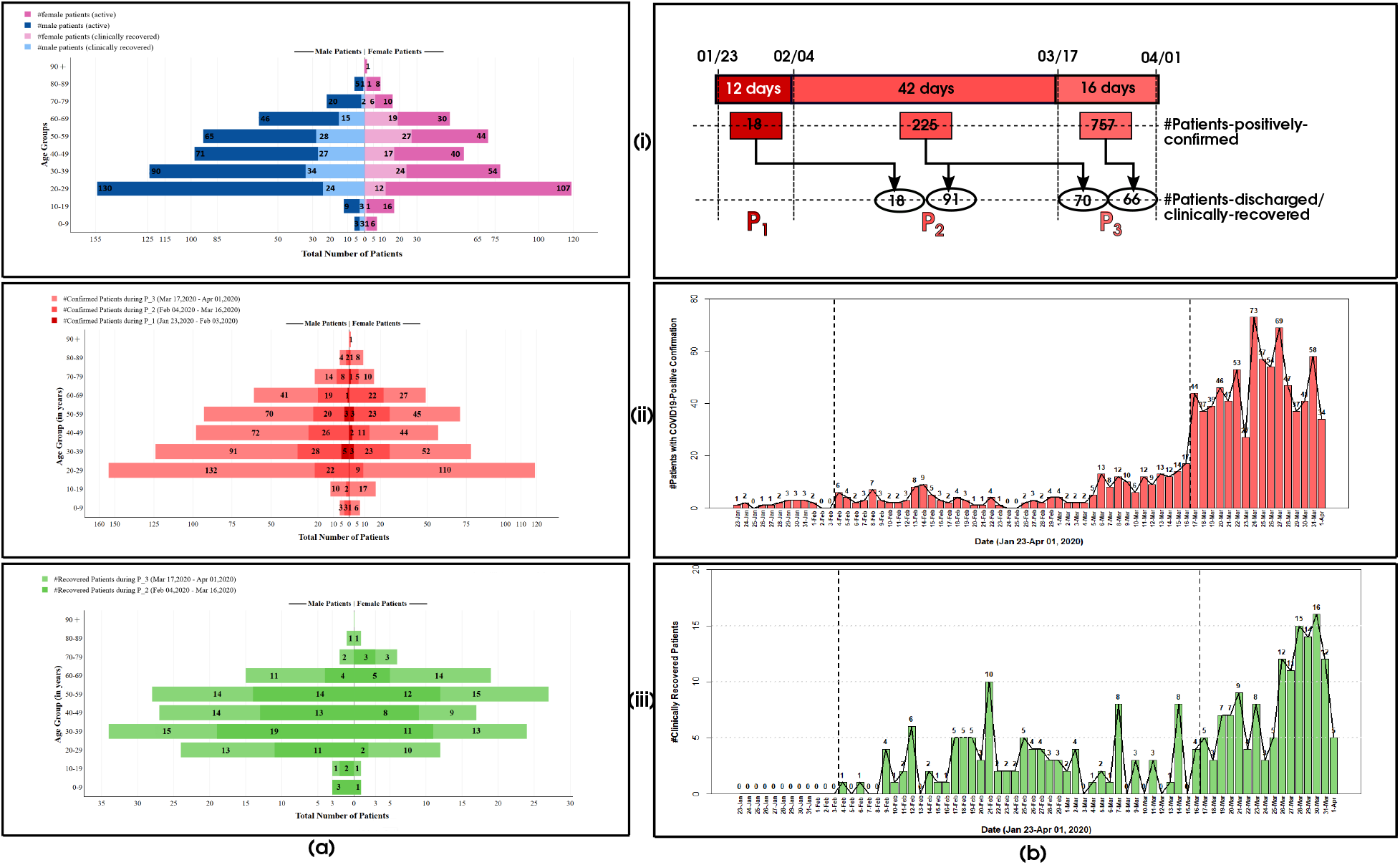
(a) Age-gender distribution observed using stacked population pyramids of COVID-19 infected and recovered, and (b) periodization of January 23-April 01, 2020, using timelines, for COVID-19 patients in Singapore, for (i) general population trends, (ii) COVID-19 positive confirmed patients, and (iii) recovered and discharged patients. The population size is (i) 1000 (576M, 424F), (ii) 1000 (576M, 424F), and (iii) 245 (137M, 108F).

**Age-gender distribution:** Using age-groups of 10 years, we observe that the age-gender distribution of the confirmed/infected, and the recovered are different (§Figure 1(a)(i)), where the gender ratios (male to female) are 1.36 and 1.27 respectively. (*The age-gender data is given as percentages in Table S1, Supplementary File*.) The highest counts of the confirmed and the recovered are in the age-groups (20-29) years (§Figure 1(a)(ii)) and (30-39) (§Figure 1(a)(iii)) years, respectively, demonstrating a conspicuous shift of the affected population to the age groups (20-39) years.

**Periodization:** The first imported and local transmission cases were confirmed on January 23 and February 4, respectively. There is a sudden spike in the daily confirmations on March 17. We periodize using these events for piecewise analysis of the pandemic progression (§Figure 1(b)). Given the cumulative counts of positive confirmed cases, *N_i_*, discharged/recovered cases, *N_r_*, and deceased, *N_d_*, we get:

1. Period *P*_1_ (*period of imported cases*) during January 23-February 3, (*N_i_*=18, *N_r_*=0, *N_d_*=0).
2. Period *P*_2_ (*period of local transmission*) during February 4-March 16, (*N_i_*=243, *N_r_*=109, *N_d_*=0).
3. Period *P*_3_ (*period of community transmission*) during March 17-April 1, (*N_i_*=1000, *N_r_* =245, *N_d_*=3).

We observe from the periodwise age-gender distribution that *P*_1_ is different from both *P*_2_ and *P*_3_ (§Figure 1(a)(ii),(a)(iii)). The ratio of recovered to infected, *r_ri_*, i.e., 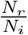, has increased from 0.00 to 0.45, again dipped to 0.245, through *P*_1_ to *P*_3_. However, since the trend in *r_ri_* does not explicitly show the influence of shifts in the age-gender distribution (§Figure 1(a)(ii)) of *N_i_* and *N_r_* on Δ*t*_LOS_, we analyse the time-varying Δ*t*_LOS_ using descriptive statistics and regression.

**LOS analysis:** Descriptive statistical analysis of Δ*t*_LOS_ in groups based on gender, age, and period give the preliminary findings of the variables on Δ*t*_LOS_. We use two appropriate regression models to estimate Δ*t*_LOS_. Firstly, we use the time-series of Δ*t*_LOS_ and fit a line using the loess model. Loess is a non-parametric local regression model for smoothening empirical time-series data [8] and scatterplots [9]. Secondly, we consider Δ*t*_LOS_ as an observed count variable and fit the frequency distribution of Δ*t*_LOS_ into a multivariate linear regression models. We use semi-parametric generalized linear models (GLM) for regression, with age, gender, and period are predictor variables, and Δ*t*_LOS_ as the dependent variable. For the period-wise analysis of Δ*t*_LOS_, we group the discharged patients based on their positive confirmation date, 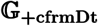. An alternative grouping is also possible based on their discharge date, 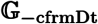. Since the protocols followed in the hospital are similar for patients with closer admission dates, the 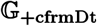 grouping shows more cohesive descriptive statistics of Δ*t*_LOS_ than the 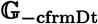 grouping. Hence, we use the 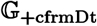 grouping here. (*The descriptive statistics of* Δ*t_LOS_ with* 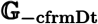 *grouping is given in Table S2, Supplementary File*.)

We use the loess model [8] to smooth the conditional means in the observations of Δ*t*_LOS_ for getting its trendline. Loess uses linear least squares regression model, *y* = *g*(*x*) + *ε*, with independent and dependent variables, *x* and *y*, respectively, a smooth function *g* (we use the Gaussian function), and a 0-mean constant-scale random variable as the error value, *ε*. The loss function *ε_w_*(*x*) = *w*(*x*).(*y* - *g*(*x*))^2^ is minimized using a weight function *w*(*x*). Loess uses a key parameter, span *α*, that is the control parameter for the size of the local neighborhood. When *α* < 1, *w*(*x*) = (1 - |*d*|^3^)^3^, the tricube weight function with d is the normalized distance from data points to the fitted curve, and otherwise, *w*(*x*) =0.

Given that school children and senior citizens are considered vulnerable groups, we implement age binning for performing hypothesis testing and regression modeling. These age-bins (0-19), (20-59), and (60-89) are referred to as *A*_1_, *A*_2_, and *A*_3_, respectively. Thus, we use the binning of {male, female}, {*P*_1_, *P*_2_, *P*_3_}, and {*A*_1_, *A*_2_, *A*_3_} for gender, period, and age, respectively, for hypothesis testing and determining appropriate GLM’s. We use the Kruskal-Wallis H (KWH) test of Δ*t*_LOS_ with respect to age-bins, and with the period, separately. The KWH test is a rank-based non-parametric test to determine if two or more groups of an independent ordinal/continuous variable (age or period) have statistically significant differences with the continuous dependent variable (Δ*t*_LOS_). We use the Mann-Whitney-Wilcoxon (MWW) test of Δ*t*_LOS_ with respect to gender. The MWW test is a non-parametric two-sample test to verify the null hypothesis that the distribution of a continuous variable (Δ*t*_LOS_), which is not normal, is the same in two or more independent groups formed based on a nominal/categorical independent variable (gender).

We determine the appropriate GLM by experimenting with Poisson (PRM) and negative binomial (NBM) distributions for Δ*t*_LOS_. These distributions are commonly used for LOS, owing to its naturally skewed distribution [10], and for over-dispersed data [11], such as LOS. Here, we fit the model using frequency distribution of LOS, and model has LOS as dependent variable *Y*, and age, gender, and period of infection as predictor variables *X*_1_, *X*_2_, and *X*_3_, respectively. They are used as numerical, nominal, and ordinal type variables, respectively. We assume the frequency distribution of *Y* to be Poisson with *E*(*Y*) = *V ar* (*Y*) = *μ*, or negative binomial, where *V ar* (*Y*) = *μ* is relaxed. Thus, the GLM with intercept *β*_0_, regression coefficients, *β_i_* for *i* = 1, 2,3, and error term *ε* is given as:

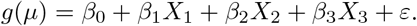

## 3 Results

**Descriptive statistical analysis:** Δ*t*_LOS_ observed during the entire period, January 23-April 01 show the descriptive statistics (§Figure 2 (i)-(iii)) with the following (#patients, mean, standard deviation, median, interquartile range), (*N*, *M*, *SD*, Mdn, IQR), values:

- Overall: (*N*=245, *M*=11.4, *SD*=5.8, Mdn=11, IQR=[7, 14]) days.
- Gender-wise (§Figure 2(i)): (*N*=108, *M*=11.28, *SD*=5.85, Mdn=11, IQR=[7.75, 14]) days for females and (*N*=137, *M*=11.42, *SD=* 5.77, Mdn=11, IQR=[7, 15]) days for males.

• Age-wise, for age-groups in years given in parentheses (§Figure 2(ii)):

- *A*_1_: (*N*=4, *M*=6.75, *SD*=7.04, Mdn=4.5, IQR=[3.25, 8]) days for (0-9), (*N*=4, *M* =13.25, *SD*=3.5, Mdn=13.5, IQR=[11.25, 15.5]) days for (10-19),
- *A*_2_: (*N*=36, *M* =10.44, *SD*=4.88, Mdn=9.5, IQR=[6, 14]) days for (20-29), (*N*=58, *M* =11.33, *SD*=5.64, Mdn=11, IQR=[8, 13]) days for (30-39), (*N*=44, *M* =10.64, *SD*=4.73, Mdn=11, IQR=[6.75, 13.25]) days for (40-49), (*N*=55, *M* =12.78, *SD*=7.21, Mdn=11, IQR=[8, 16.5]) days for (50-59),
- *A*_3_: (*N*=34, *M* =12, *SD*=5.53, Mdn=11.5, IQR=[9, 14.75]) days for (60-69), (*N*=8, *M*=9.38, *SD*= 5.55, Mdn=9, IQR=[6.25, 13.25]) days for (70-79), (*N*=2, *M*=7.5, *SD*=4.95, Mdn=7.5, IQR=[5.75, 9.25] days for (80-89).

• Period-wise, based on 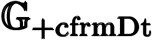 grouping (§Figure 2(iii)):

- (N =18, *M* =17.61, *SD*=6.6, Mdn=17, IQR=[11.5, 22.75]) days for *P*_1_,
- (N =161, *M* =11.88, *SD*=5.81, Mdn=12, IQR=[8, 16]) days for *P*_2_,
- (*N*=66, *M*=8.58, *SD*=3.35, Mdn=9, IQR=[6, 11]) days for *P*_3_.

**Figure 2:**
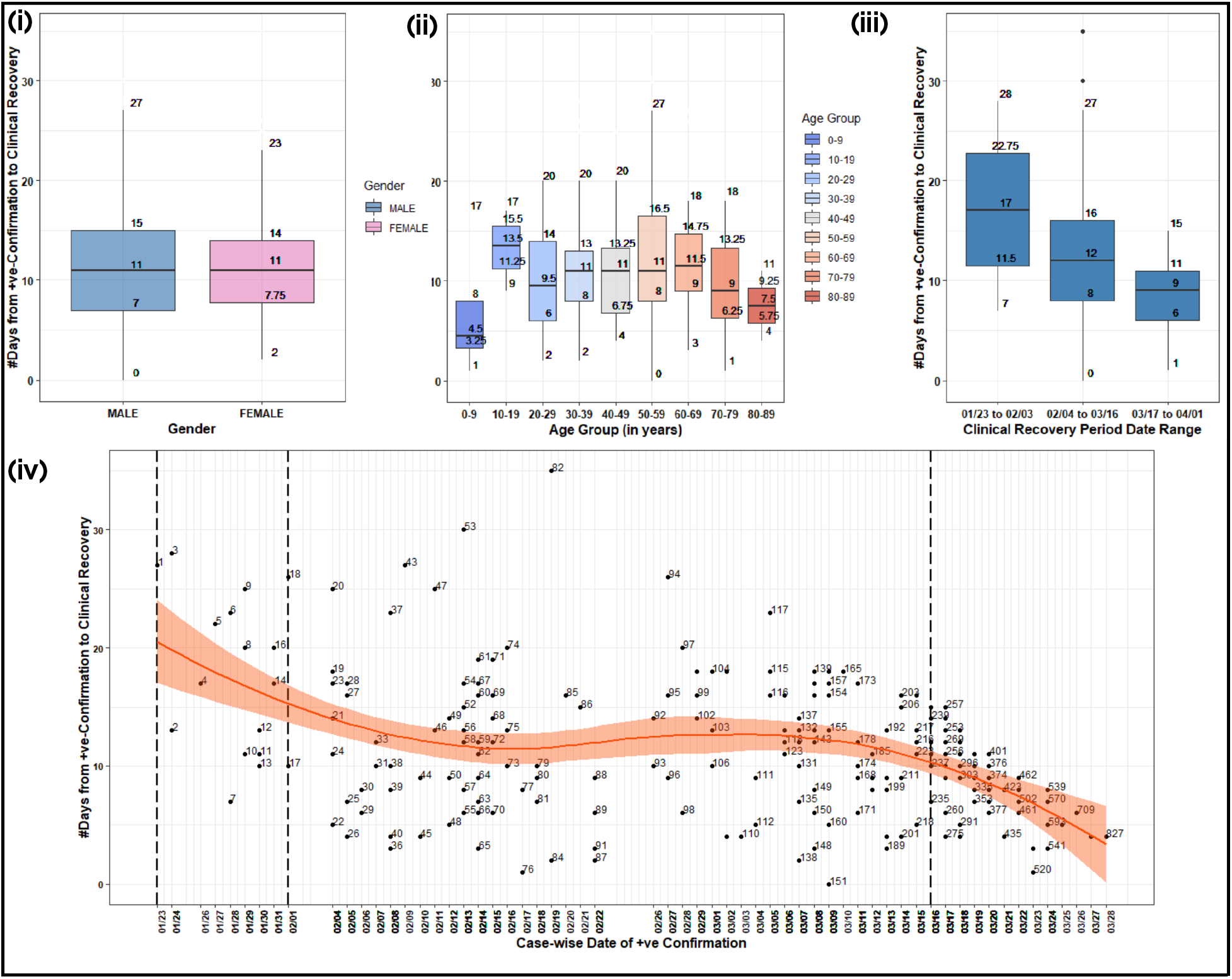
Box-and-whisker plots of LOS of 245 discharged COVID-19 patients during January 23-April 01, 2020, in Singapore, based on (i) gender, (ii) age, and (iii) period, with 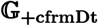 grouping. (iv) Scatter plot of the length of in-hospital stay for 245 discharged patients with the timeline of their COVID-19 positive confirmation, with a trend-line estimated using a loess model. The fitted loess curve and its error ribbon show a decrease in LOS during the period.

We observe that the Δ*t*_LOS_ is overall lesser in this dataset than reported in early analyses of COVID-19 patients, i.e., 15 days [6] and 20 days [7]. The overall median of Δ*t*_LOS_ and the gender-weighted median is 11 days, while the age-weighted and period-weighted medians are 10.7 and 11.6 days, respectively. Thus, the overall descriptive statistical analysis gives a conservative estimate of Δ*t*_LOS_ to be ~11 days.

There is a monotonous decrease in Δ*t*_LOS_ with time, demonstrated from *P*_1_ to *P*_3_. To analyse this observation further, we look at the period-wise descriptive statistics for Δ*t*_LOS_ with respect to gender, using 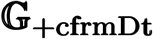 grouping.

- For females: (*N*=9, *M* =16.33, *SD*=6.44, Mdn=13, IQR=[11, 22]) days for *P*_1_, (*N*=69, *M* =11.8, *SD*=6.1, Mdn=12, IQR=[7, 15]) days for *P*_2_, and (*N*=30, *M*=8.57, *SD*=3.36, Mdn=9, IQR=[6.25, 11]) days for *P*_3_.
- For males: (*N*=9, *M* =18.89, *SD*=6.88, Mdn=20, IQR=[17, 23]) days for *P*_1_, (*N*=92, *M* =11.93, *SD*=5.61, Mdn=12, IQR=[8, 16]) days for *P*_2_, and (*N*=36, *M*=8.22, *SD*=3.37, Mdn=8, IQR=[5.75, 11]) for *P*_3_.

We observe that Δ*t*_LOS_ has similar descriptive statistics across both genders in *P*_3_ than in *P*_1_ or *P*_2_. The IQR of Δ*t*_LOS_ decreases from *P*_1_ to *P*_3_ for both genders, indicating the decrease in the *measure of the spread* of Δ*t*_LOS_ with time. (*The age-groupwise breakdown of descriptive statistics is given in Table S2, with corresponding box-and-whisker plots in Figure S2 in the Supplementary File*.)

There is a strong influence of age as well as period on Δ*t*_LOS_, unlike gender. Δ*t*_LOS_ is relatively lower for the age group of (20-29) years, specifically. This result is significant as the age group of (20-29) years contributes the highest (27.3%) to the infected population and relatively high (14.6%) to the recovered population. At the same time, the number of COVID-19 cases has increased in this age group over time, as 88.6% of the patients in this age group have been confirmed positive in *P*_3_. Thus, our key conclusion is that since the most susceptible group of people has lower Δ*t*_LOS_, the overall Δ*t*_LOS_ is influenced by age to decrease with time progressively.

**Loess model:** We smooth the time-series of the Δ*t*_LOS_, represented as a scatter plot based on hospital admission dates (§Figures 2 (iv)) using loess^7^ implemented using loess with default span *α* = 0.75 in R [12]. The loess model for discharged patients during January 23-April 01 gives the following degrees of freedom (*df*) and residual standard error (RSE) for *N* patients: (*N*=245, *df*=5.37, RSE=145.4) (§Figure 2(iv)). Δ*t*_LOS_ has a slope of −25° in *P*_3_. Thus, the key conclusion from the local regression model on the time-series is the negative slope, i.e., a *downward* trend in Δ*t*_LOS_ in *P*_3_. (*Smoothing of periodwise observations using separate loess models is given in Figure S1 in Supplementary File*.)

**Multivariate linear regression model:** The regression model can be made piecewise with respect to a predictor variable in the case of non-monotonous behavior of the observed variable with respect to the predictor variable. We observe that the median Δ*t*_LOS_ monotonously increases from age-bins (20-29) to (50-59), and then further monotonously decreases with age (§Figure 2 (ii)). There is anomalous behavior in age-bin (0-19). Unlike age, we observe that Δ*t*_LOS_ shows monotonous behavior in the case of gender and period (§Figure 2 (i), (iii)). Hence, we use the piecewise GLMs for regression with respect to age-bins.

We have implemented the hypothetical tests using kruskal.test and wilcox.test in Stats package in R [12]. The null hypothesis H_0_ is that “all groups are statistically similar,” and the alternative *H*_1_ hypothesis is that “all groups are not statistically similar.” We reject the *H*_0_ when *p*-value <5%. Our results (§Table 1) show that groups based on the period with 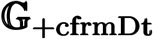 grouping show statistically significant differences, thus indicating that period influences Δ*t*_LOS_.

**Table 1:**
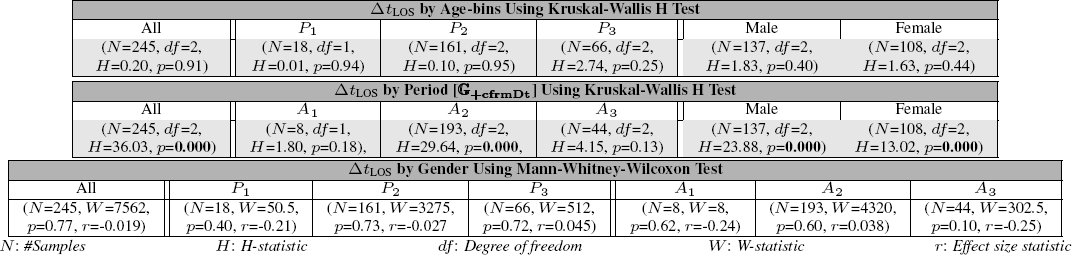
Hypothesis testing results of 245 discharged patients using the Kruskal-Wallis H test of Δ*t*_LOS_ with respect to age and period, and using the Mann-Whitney-Wilcoxon test of Δ*t*_LOS_ with respect to gender.

With respect to our study on vulnerable populations, we see that *A*_2_ sub-population with the 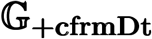 grouping shows statistical significance with respect to the period, unlike *A*_1_ and *A*_3_. We observe that both male and female separately have statistical differences with respect to period. Overall, these results confirm the use of age-binning for piecewise GLMs.

We consider two modeling scenarios for GLM, where we use age, gender, and period as independent variables: *M*_1_ which is a single GLM for all 245 patients; and *M*_2_ with three piecewise linear GLM’s for *A*_1_, *A*_2_, and *A*_3_, respectively. (*The count distribution and approximation of distribution functions are shown in Figure S3, Supplementary File*.) We have implemented these models using the glm in Stats package in R [12]. For GLM with Poisson and binomial families, the dispersion is fixed at 1.0, and the number of parameters (*k*) is the same as the number of coefficients in the regression model [12]. The negative binomial distribution has an additional parameter to model over-dispersion in the data. We use the Akaike Information Criterion (AIC) and its corrected version for a small sample size (AIC_c_) for determining the *goodness of fit* of our proposed models. For the number of samples (N) in the data, we use AIC when 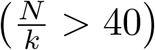, and AIC_c_, otherwise [13]. We infer the following from our results (§Table 2):

- The “period” variable is significant for both models and all scenarios, except *M*_2_ (age:(0-19)), with *p*-value <5% for corresponding coefficients.
- The “age” and “gender” variables are significant only in the PRM for *M*_2_ (age:(20-59)) and *M*_2_ (age:(60-89)) respectively, with *p*-value of corresponding coefficients being <5%, confirming the age-bin (20-59) getting progressively affected with time, and co-morbidities in older males, respectively.
- The NBM shows lower medians and the variances (observable from range and IQR) of deviance residuals, and AIC/ AIC_c_ and more consistent Δ*t*_LOS_ for maximum likelihood value than the PRM. Thus, NBM is a better fit than PRM.
- Overall, the small sample size in *M*_2_ (age:(0-19)) makes the modeling erroneous. For both PRM and NBM, the remaining piecewise models in scenario *M*_2_ are a better fit than the single model, *M*_1_, owing to the lower error measures (AIC/ AIC_c_ and deviance residuals) as well as the consistency in the maximum likelihood value. We also find that the errors, MSE and RMSE in *M*_1_, are reduced when we exclude the samples of (age:(0-19)) from the observations.

**Table 2:**
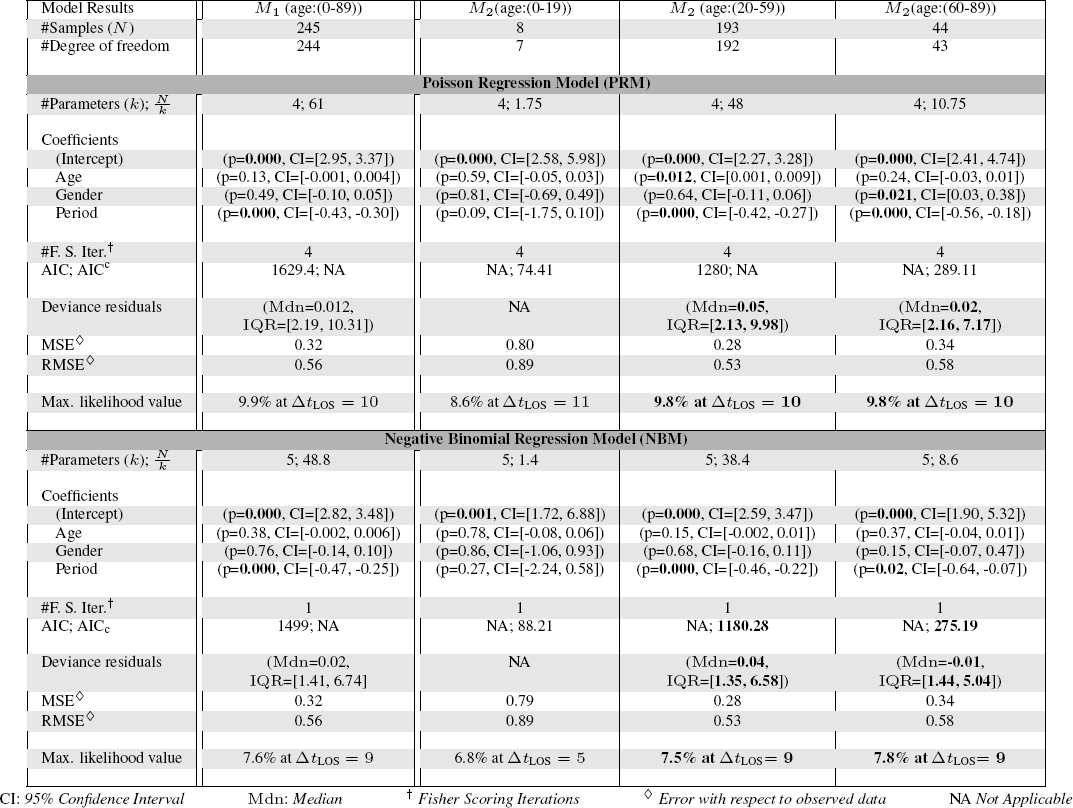
Results of the generalized linear models with age, gender, and period as independent variables, using Poisson distribution and negative binomial distribution of Δ*t*_LOS_ with 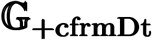 grouping.

Overall, choosing NBM and *M*_2_, we estimate Δ*t*_LOS_ = 9 days with a maximum likelihood of 7.5%. Since we do not have a large number of variables to discard, we retain the “gender” and “age” variables in the model despite its relative insignificance compared to “period.” Our additional experiments show that the error values increase in the absence of the period as an independent variable. The key conclusion from the hypothesis testing and multivariate regression analysis is that a GLM with NBM for *M*_2_ is the best model to estimate Δ*t*_LOS_.

## 4 Discussion

In summary, Δ*t*_LOS_ can be estimated to have decreased to 9 days during January 23-April 01, 2020, in Singapore. We have shown that the recovery in the vulnerable groups, such as the children and the older population, has become insignificant with time. Our work shows that this temporal shift in the demographic structure has led to the overall decrease in Δ*t*_LOS_, which is reflected in two results. Firstly, we observe “period” causes significant differences in the distribution of Δ*t*_LOS_ in both the entire population, as well as, specifically, in age-bin (20-59) (A_2_) subpopulation. Secondly, we find that “age” is a significant variable for the multivariate regression model with PRM for the population of the age-bin (20-59) years in *M*_2_. These indicate that our age-binning and periodization are significant in analyzing and estimating Δ*t*_LOS_. Periodwise piecewise GLM’s of observations in *A*_2_, with age and gender as predictor variables, show that the estimate of Δ*t*_LOS_ reduces from ~16 days in *P*_1_ to ~9 days in *P*_2_, and further to ~8 days in *P*_3_. (*Details of the periodwise GLMs are in Table S3 in Supplementary File*.) Thus, our findings of the progressive decrease in Δ*t*_LOS_ are confirmed using these models. The overall reduction in Δ*t*_LOS_ is an outcome of the significance of Δ*t*_LOS_ in *A*_2_, which is the non-vulnerable population.

Our findings can be attributed to the early response of restricted mobility of the vulnerable groups. The vulnerable population becoming insignificant can be an outcome of the increase in positive confirmations in the non-vulnerable group. However, our work has the limitation of modeling recovery exclusively, without the interaction with the distribution of daily positive confirmations. With N_i_ = 1000 in Singapore by April 1, but *N_r_* = 245, we have not analyzed the outcome of the remaining 755 patients. Our study can be enriched by doing a similar analysis on all the cases confirmed by April 1, closed by recovery and death. Our regression model can be improved to include interaction effects, which could be effective if we have additional data on other variables, e.g., co-morbidities, whose data is not available in the public domain. Nevertheless, our estimated Δ*t*_LOS_ based on the first 245 discharged patients is useful for assessing the hospital load in terms of required resources for the in-hospital stay of the COVID-19 cases.

## Data Availability

The data used for this study is curated from the press releases published online by the Ministry of Health, Government of Singapore, and is available at https://github.com/vrrani/COVID19-Singapore

## Acknowledgements

The authors would like to thank IIIT Bangalore, and particularly, the Graphics-Visualization-Computing Lab and the E-Health Research Center, for supporting this work. The authors appreciate the constructive review from Srujana Merugu in shaping this work. This work has been partially supported by the IBM Shared University Grant awarded to J. Sreevalsan-Nair and the Visvesvaraya Ph.D. Fellowship by the Ministry of Electronics and IT (MeitY), Government of India, awarded to R. R. Vangimalla.

## Footnotes

2 https://www.who.int/dg/speeches

3 https://www.who.int/emergencies/diseases/novel-coronavirus-2019/situation-reports

4 This is based on the unofficial counts are, as retrieved on April 2, 2020, from https://www.worldometers.info/coronavirus/coronavirus-cases

5 https://www.cdc.go.kr/board/board.es?mid=a30402000000&bid=0030&tag=&act=view&list_no=366352

6 https://www.moh.gov.sg/news-highlights/

7 The loess model is a default local regression model used for a sample with less than 1000 observations in Stats package in R.

---

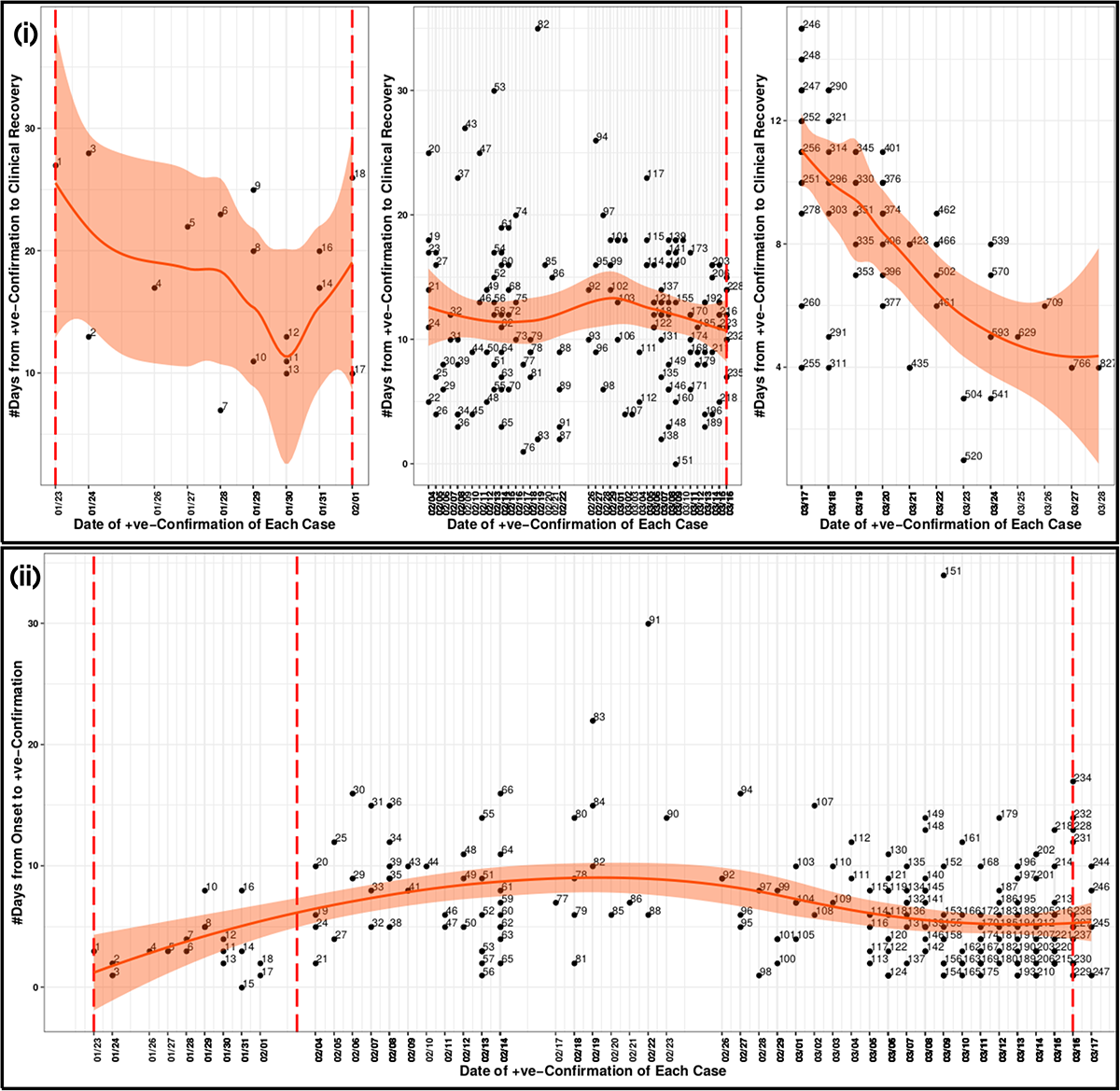

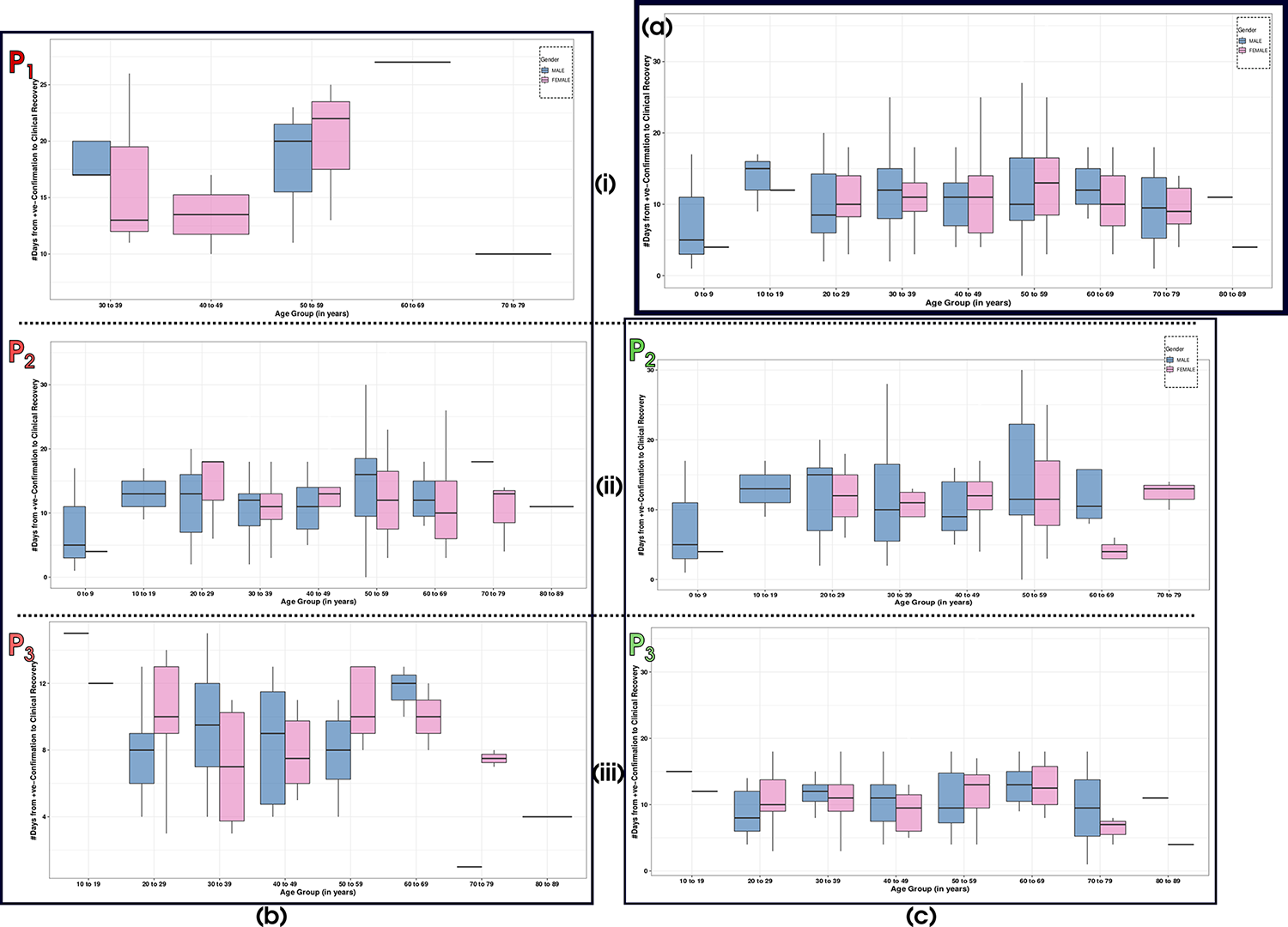

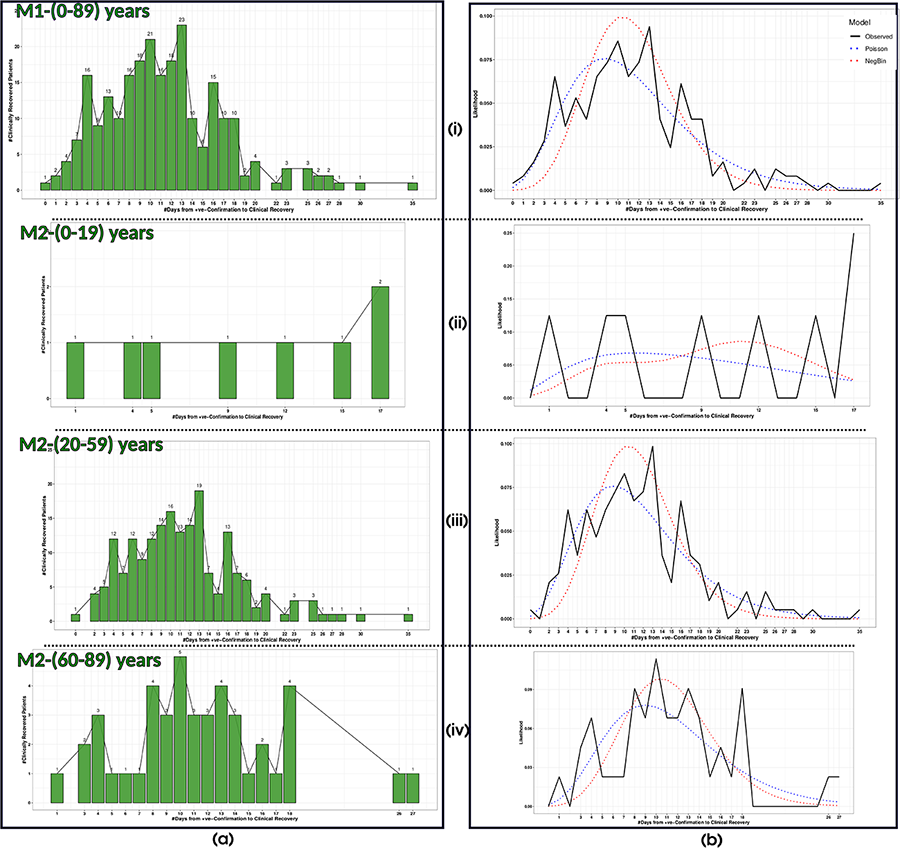

## References

[1] Rachael Pung, Calvin J. Chiew, Barnaby E. Young, Sarah Chin, Mark I-C Chen, Hannah E. Clapham, Alex R. Cook, Sebastian Maurer-Stroh, Matthias P. H. S. Toh, Cuiqin Poh, Mabel Low, Joshua Lum, Valerie T. J. Koh, Tze M. Mak, Lin Cui, Raymond V. T. P. Lin, Derrick Heng, Yee-Sin Leo, David C. Lye, and Vernon J. M. Lee. Investigation of three clusters of COVID-19 in Singapore: implications for surveillance and response measures. The Lancet, 395:1039–46, 2020.

[2] Nanshan Chen, Min Zhou, Xuan Dong, Jieming Qu, Fengyun Gong, Yang Han, Yang Qiu, Jingli Wang, Ying Liu, Yuan Wei, and et al. Epidemiological and clinical characteristics of 99 cases of 2019 novel coronavirus pneumonia in Wuhan, China: a descriptive study. The Lancet, 395(10223):507–513, 2020.

[3] Xiaobo Yang, Yuan Yu, Jiqian Xu, Huaqing Shu, Jia’an Xia, Hong Liu, Yongran Wu, Lu Zhang, Zhui Yu, Minghao Fang, Ting Yu, Yaxin Wang, Shangwen Pan, Xiaojing Zou, Shiying Yuan, and You Shang. Clinical course and outcomes of critically ill patients with SARS-CoV-2 pneumonia in Wuhan, China: a single-centered, retrospective, observational study. The Lancet Respiratory Medicine, 2020.

[4] Adam J Kucharski, Timothy W. Russell, Charlie Diamond, Yang Liu, John Edmunds, Sebastian Funk, and Rosalind M. Eggo. Early dynamics of transmission and control of COVID-19: a mathematical modelling study. The Lancet Infectious Diseases, 2020.

[5] Qun Li, Xuhua Guan, Peng Wu, Xiaoye Wang, Lei Zhou, Yeqing Tong, Ruiqi Ren, Kathy S.M. Leung, Eric H.Y. Lau, Jessica Y. Wong, Xuesen Xing, Nijuan Xiang, Yang Wu, Chao Li, Qi Chen, Dan Li, Tian Liu, Jing Zhao, Man Liu, Wenxiao Tu, Chuding Chen, Lianmei Jin, Rui Yang, Qi Wang, Suhua Zhou, Rui Wang, Hui Liu, Yinbo Luo, Yuan Liu, Ge Shao, Huan Li, Zhongfa Tao, Yang Yang, Zhiqiang Deng, Boxi Liu, Zhitao Ma, Yanping Zhang, Guoqing Shi, Tommy T.Y. Lam, Joseph T. Wu, George F. Gao, Benjamin J. Cowling, Bo Yang, Gabriel M. Leung, and Zijian Feng. Early Transmission Dynamics in Wuhan, China, of Novel Coronavirus-Infected Pneumonia. New England Journal of Medicine, 382(13):1199–1207, 2020.

[6] Cleo Anastassopoulou, Lucia Russo, Athanasios Tsakris, and Constantinos Siettos. Data-based analysis, modelling and forecasting of the COVID-19 outbreak. PloS one, 15(3):e0230405, 2020.

[7] Fei Zhou, Ting Yu, Ronghui Du, Guohui Fan, Ying Liu, Zhibo Liu, Jie Xiang, Yeming Wang, Bin Song, Xiaoying Gu, Lulu Guan, Yuan Wei, Hui Li, Xudong Wu, Jiuyang Xu, Shengjin Tu, Yi Zhang, Hua Chen, and Bin Cao. Clinical course and risk factors for mortality of adult inpatients with COVID-19 in Wuhan, China: a retrospective cohort study. The Lancet, 395(10229):1054–62, 2020.

[8] William S Cleveland and Susan J Devlin. Locally weighted regression: an approach to regression analysis by local fitting. Journal of the American statistical association, 83(403):596–610, 1988.

[9] William G Jacoby. Loess:: a nonparametric, graphical tool for depicting relationships between variables. Electoral Studies, 19(4):577–613, 2000.

[10] Evelene M Carter and Henry WW Potts. Predicting length of stay from an electronic patient record system: a primary total knee replacement example. BMC medical informatics and decision making, 14(1):26, 2014.

[11] Liming Xiang, Andy H Lee, Kelvin KW Yau, and Geoffrey J McLachlan. A score test for overdispersion in zero-inflated poisson mixed regression model. Statistics in medicine, 26(7):1608–1622, 2007.

[12] R Core Team. R: A Language and Environment for Statistical Computing. R Foundation for Statistical Computing, Vienna, Austria, 2013. ISBN 3-900051-07-0.

[13] K. P. Burnham and D. R. Anderson. Model selection and inference: a practical information-theoretic approach. Springer Verlag, 1998.

